# Analysis and prediction of Covid-19 spreading through Bayesian modelling with a case study of Uttar Pradesh, India

**DOI:** 10.1101/2020.08.25.20180265

**Authors:** Deepmala, Nishant Kumar Srivastava, Vineet Kumar, Sanjay Kumar Singh

## Abstract

The pandemic of coronavirus disease 2019 (COVID-19) started in Wuhan, China, and spread worldwide. In India, COVID-19 cases increased rapidly throughout India. Various measures like awareness program, social distancing, and contact tracing have been implemented to control the COVID-19 outbreak. In the absence of any vaccine, the prediction of the confirmed, deceased, and recovered cases is required to enhance the health care system’s capacity and control the transmission. In this study, the cumulative and daily confirmed, deceased, and recovered cases in Uttar Pradesh, India, were analyzed. We used the logistic and Gompertz non-linear regression model using a Bayesian paradigm. We build the prior distribution of the model using information obtained from some other states of India, which are already reached at the advanced stage of COVID-19. Results from the analysis indicated that the predicted maximum number of confirmed, deceased, and recovered cases will be around 1157335, 5843, and 1145829 respectively. The daily number of confirmed, deceased, and recovered cases will be maximum at 104th day, 73rd day, and 124th day from 16 June 2020. Further from this analysis we can conclude that the COVID-19 will be over probably by early-June, 2021. The analysis did not consider any changes in government control measures. We hope this study can provide some relevant information to the government and health officials.

## 1. Introduction

The world is facing the outbreak of the coronavirus which is a highly contagious disease also it is declared as a global public health emergency by the World Health Organisation (WHO) [1]. It is originated in Wuhan, Hubei, China in December 2019 and spread all over the world. The World Health Organization officially named the disease COVID-19 and classified the COVID-19 outbreak as pandemic on March 11, 2020 [9, 21] This virus is highly infectious and transmittable. Precautionary measures for COVID-19 contain maintaining social distancing, wearing masks, cleaning hands regularly, avoiding touching the eyes, nose, and mouth (WHO, 2020). The first confirmed case of COVID-19 in India was recorded on 30 January 2020. Since then, the number of confirmed cases and deceased cases is continuously increasing and most of the states affected by this virus COVID-19. This disease has recorded millions of cases and thousands of deaths due to rapid pandemic potential. At present, India is the most affected country in Asia and has the third-largest number of confirmed cases in the world after the USA and Brazil [11]. It could be soon in the top spot in terms of cases [7, 22]. In India, Maharastra has the highest number of cases of COVID-19 [6]. Currently, in Maharastra, more than four lakhs cases are recorded. Tamil Nadu, Andhra Pradesh, Karnataka, NCT of Delhi and Uttar Pradesh have reported more than one lakh cases [6]. There is no vaccine until now of this virus. The absence of a vaccine creates the situation worse for the already overstretched health care system. For example, the number of hospital beds per thousand population is less than one [10]. It is indicating the miserable situation of India’s health care system.

In India, Uttar Pradesh (UP) is a state that holds a significant value concerning the economy, trade, manufacturing, and services. It has the second-largest economy in India after Maharashtra in terms of net state domestic product. As of 06 August 2020 in India, 2025423 confirmed cases had been reported. Of these, 108974 confirmed cases are from the state of Uttar Pradesh, India [6]. Uttar Pradesh, India’s largest democracy with a population of 200 million, will have difficulty controlling the transmission of COVID-19 among its population. In such a grim scenario, many outcomes are potential interest to policymakers; for example: how many confirmed, deceased, and recovered cases will be seen, and by what time? How many will be admitted to ICU? How many will need ventilators? When will be started the declining trend of COVID-19? The government could take the optimal steps to improve the health system to slow down the transmission rate by knowing even roughly the answer to these questions. Multiple strategies would be highly necessary to handle the current outbreak; these include computational modeling, statistical tools, and quantitative analyses to control the spread and the rapid development of a new treatment. It is imperative that the trends and predictions for the state of Uttar Pradesh must be studied well so that effective strategies can be implemented. We focus on the number of confirmed, deceased, and recovered cases as our target of prediction due to limited available data on the other outcomes.

Several mathematical and statistical models can be established to study and analyze the transmission process of COVID-19. So that we can accurately predict the prevalence and explore the situation of the probability of cases and the recovery or deaths. Therefore, to reduce the harm of the COVID-19 outbreak, the analysis and research of Covid-19 prediction models have become a hot research topic.

The most commonly used models for predicting infectious diseases are internet-based infectious disease prediction models, time series prediction models, and differential equation prediction models based on dynamics. Several models have been proposed in literature [5, 8, 13, 16]. The studies are very less for the India region, and some work has been done by [12, 18–20]. From an analysis point of view, non-linear models are also important methods to deal with the problem of prediction [3, 14, 15]. As we know, the mechanisms of COVID-19 spreading are not completely understood. Therefore Bayesian analysis of the epidemic spreading can be interesting. This approach has the advantage that it includes some other prior information in the model other than the data.

This study focuses on the analysis and the prediction of the epidemic situation of COVID-19 in the state of Uttar Pradesh, India, using logistic and Gompertz nonlinear regression model, which are accord with the statistical law of epidemiology. The Bayesian approach [4] is used for estimation purposes. Here prior information is obtained from other states of India.

## 2. Data

In this paper, we used the following major databases:

1. Github-Covid-19 in India available from https://github.com/imdevskp/covid-19-india-data
2. Covid19India is a open source database for India available from: https://www.covid19india.org/

## 3. Material & Methods

The main objective of the study is to make predictions of the cumulative and daily confirmed, deceased, and recovered cases in the state of Uttar Pradesh, India. For this purpose, we use Bayesian non-linear logistic model (LM) and Gompertz model (GM). Prior information is incorporated from results obtained from the fitting of classical non-linear regression model to the other five states, which are already reached to the advanced stage of the Covid-19 outbreak. These states have significant variations from each other.

Initially, we fitted logistic and Gompertz non-linear regression model to the data of cumulative cases of COVID-19 for five states: Andhra Pradesh (AP), Delhi (DL), Karnataka (KA), Maharashtra (MH), and Tamil Nadu (TN). The logistic and Gompertz non-linear regression model can be mathematically expressed as:

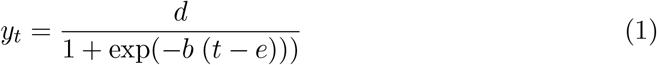

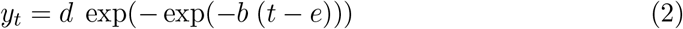

respectively. Both models have the same meaning of the parameters. Here *y_t_* represents cumulative cases (confirmed, deceased or recovered cases) of COVID-19 observed at *t^th^* time, d is the asymptote that represents the predicted maximum of cumulative cases at the end of the outbreak, b is the slope around the inflection point that represents the growth rate coefficient, e is the time at inflection that represents the time when will occur the maximum daily cases. The difference between these two models is that the logistic model is symmetric around the inflection point while Gompertz is not.

Non-linear regression models were fitted using the drm function of R package drc [17] of R statistical software. The criteria AIC (Akaike Information Criterion) and WAIC (Watanabe Akaike information criterion) are used for comparison of classical and Bayesian models, respectively. The regression coefficient (*R*^2^) is used to measure the fitting ability of various models. It is defined as

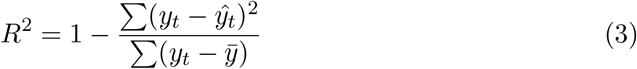

where *y_t_* is the actual cumulative confirmed, deceased, or recovered COVID-19 cases, *ȳ* is the average of the actual cumulative confirmed, deceased, or recovered COVID-19 cases. The fitting coefficient is closer to 1 that represents more accurate prediction. By using the results of the non-linear models fitted by least square estimation (LSE), we define the prior distribution of the parameters of the Bayesian non-linear models for estimating and predicting the cumulative and the daily confirmed, deceased, and recovered cases of Uttar Pradesh state. From the scatter plot of UP states and other states, we found that the parameter d that represents asymptote of the curve has a large number of variations for confirmed, deceased, and recovered cases. But we did not find such variations for the other two parameters b and e. Therefore for defining the prior distribution of parameter d, we use least square estimates and standard error of these estimates along with other important factors associated with that considered states, due to which such variations are being found. The considered important factors as additional information are: the total number of tests conducted and the total number of confirmed cases (as of till 06 August 2020). But for parameters b and e, we use only least square estimates and standard error of these estimates to specify the prior distribution.

The prior information on the parameter d can be provided as:

For confirmed cases,

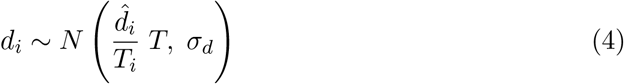

For recovered and deceased cases,

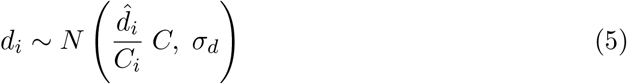

The prior information on the parameters b and e can be provided as:

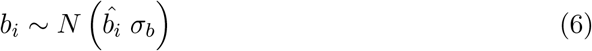

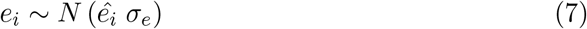

where 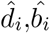 and *ê_i_* are least square estimates for *i^th^* state. *T_i_* and *C_i_* be the total number of tests conducted at i-th state and total number of confirmed cases at i-th state respectively. *T* and *C* be the total number of tests conducted in UP state and total number of cases occured in UP state respectively. *σ_d_*, *σ_b_* and *σ_e_* are standard errors of least square estimates of *d*, *b* and *e*. Parameter inference was done via the Gibbs sampling algorithm. We use Watanabe-Akaike information criterion (WAIC) and *R*^2^ to compare models. These Bayesian analyses were done using the bmrs [2] R package. The prediction on daily cases for UP state was done by using the results of Bayesian non-linear regression model for cumulative cases.

## 4. Results

In Figure 1, we plotted the bar graphs for daily confirmed cases, daily deceased cases and daily recovered cases, and the scatter plot for the cumulative confirmed cases, cumulative deceased cases, and cumulative recovered cases of COVID-19 for some states of India. Figure 1 shows that there are fluctuations in the daily confirmed cases, daily deceased cases, and the daily recovered cases of COVID-19 for all the considered states. Tables 1, 2 and 3 represent the least square estimates for the parameters of the logistic and Gompertz models with respect to considered variables. We used the regression coefficient *R*^2^ to analyze the fitting ability of the models. The value of *R*^2^ closer to one indicates a better prediction. Tables 1 and 2 show that the upper asymptote value is found to be greater through the Gompertz model as compared to those obtained using the logistic model. From Table 3, we observe that except Andhra Pradesh (AP), Gompertz models provided the greater value of asymptote than the logistic model. We see that there is a difference between the values of time inflection for both the models. The value of *R*^2^ indicates that both the models fit the data well with respect to all the considered variables. From the graphs and tables, we see that Delhi is in the controlled stage of the pandemic. Delhi crossed the inflection point of the curve and reached it’s a plateau, and there are fewer chances of fluctuation in the upper asymptote, which explains the low-value standard error of the estimate of the asymptote. From the COVID-19 updates of India, we observe that in Delhi, the death rate is about 2 %, and the recovered percentage is more than 90 %. The possible reasons behind this achievement may be the vast screening of COVID-19, effective containment, patients treatment, and contact tracing. Maharastra has the highest number of cumulative confirmed cases and the cumulative deceased cases (as of till 06 August 2020). Except Delhi the COVID-19 pandemic is not in the plateau for other considered states, so there is much uncertainty regarding the asymptote of the curve, which describes the high values of the standard error of the estimate of the asymptote. AIC values from Tables 1 and 3 showed that the logistic model was the best model for the data of the majority of the states. Table 2 showed that the logistic model was the best model only to the data of Andhra Pradesh and Karnataka, and for the rest of the states, the Gompertz model was found be best.

**Figure 1.**
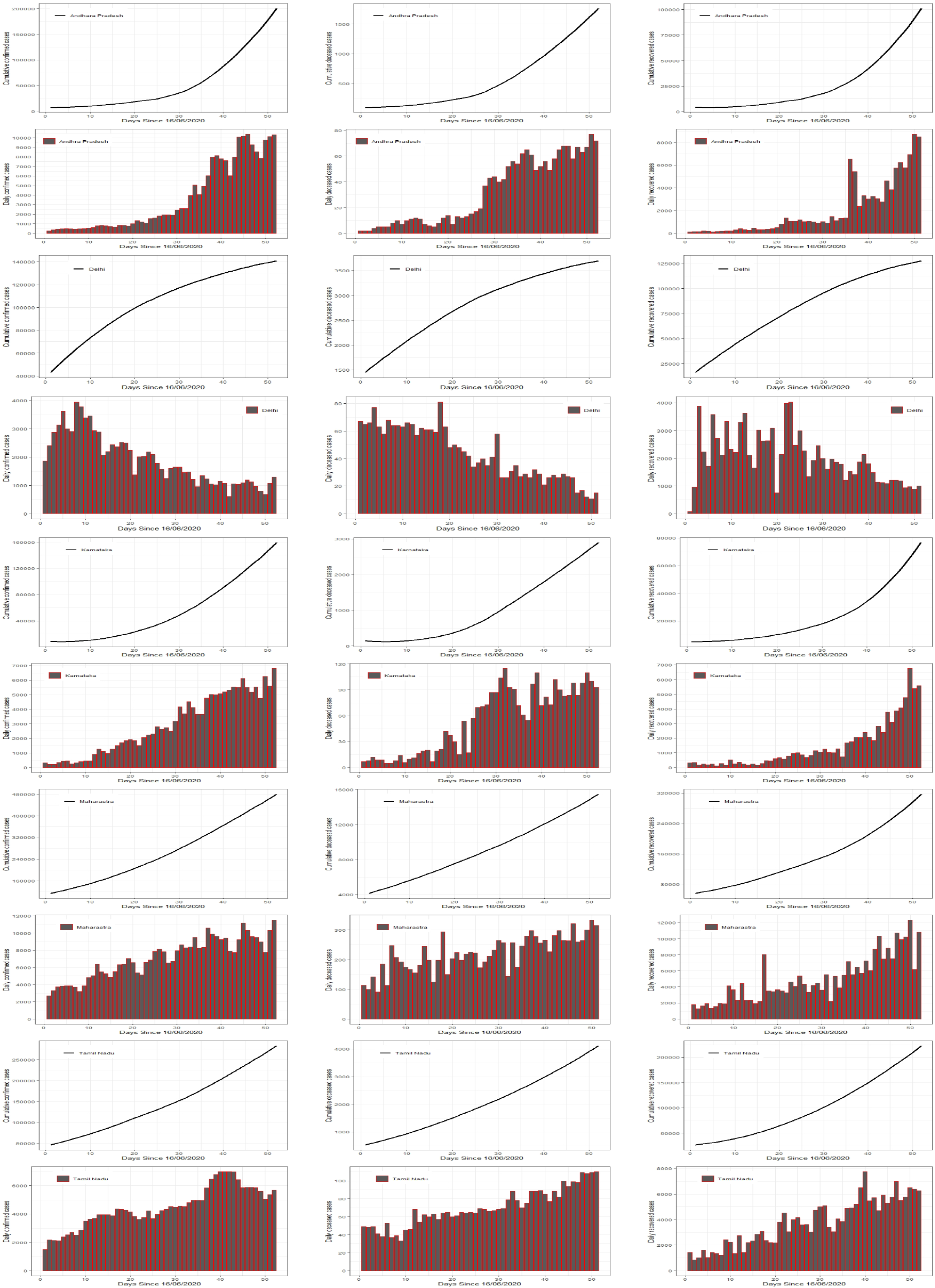
Bar plots for daily confirmed, deceased and recovered cases, and scatter plots for cumulative confirmed, deceased and recovered cases of the Covid-19 for each state analyzed

**Table 1.** Least square estimates and their respective standard errors of the parameters, *R*^2^ and AIC for the models fitted to the cumulative confirmed cases of Covid-19 of the considered states.

| Model | State | b | SE | d | SE | e | SE | R2 | AIC |
| --- | --- | --- | --- | --- | --- | --- | --- | --- | --- |
| LM | AP | 0.0869 | 0.0033 | 719391 | 153346 | 63 | 4 | 0.9865 | 1001 |
| GM |  | 0.0194 | 0.0002 | 4139788 | 292531 | 109 | 1 | 0.9326 | 1036 |
| LM | DL | 0.0781 | 0.0018 | 143090 | 958 | 10 | 0 | 0.7329 | 925 |
| GM |  | 0.0560 | 0.0009 | 148970 | 763 | 4 | 0 | 0.7374 | 871 |
| LM | KA | 0.0832 | 0.0008 | 278629 | 5560 | 49 | 0 | 0.9978 | 852 |
| GM |  | 0.0190 | 0.0002 | 1573939 | 35590 | 95 | 1 | 0.9985 | 922 |
| LM | MH | 0.0383 | 0.0003 | 1082128 | 23400 | 58 | 1 | 0.9621 | 921 |
| GM |  | 0.0095 | 0.0000 | 5261002 | 30264 | 143 | 1 | 0.9626 | 958 |
| LM | TN | 0.0481 | 0.0009 | 492941 | 18999 | 47 | 1 | 0.9607 | 968 |
| GM |  | 0.0151 | 0.0001 | 1286628 | 22769 | 80 | 1 | 0.9601 | 962 |

**Table 2.** Least square estimates and their respective standard errors of the parameters, *R*^2^ and AIC for the models fitted to the cumulative deceased cases of Covid-19 of the considered states.

| Model | State | b | SE | d | SE | e | SE | R2 | AIC |
| --- | --- | --- | --- | --- | --- | --- | --- | --- | --- |
| LM | AP | 0.0840 | 0.0029 | 3445 | 288 | 51 | 2 | 0.9967 | 510 |
| GM |  | 0.0186 | 0.0002 | 21967 | 1151 | 102 | 1 | 0.9947 | 535 |
| LM | DL | 0.0663 | 0.0007 | 3867 | 12 | 8 | 0 | 0.9994 | 447 |
| GM |  | 0.0475 | 0.0005 | 4044 | 13 | 2 | 0 | 0.9997 | 417 |
| LM | KA | 0.1044 | 0.0025 | 3679 | 96 | 40 | 1 | 0.9890 | 539 |
| GM |  | 0.0359 | 0.0022 | 7347 | 683 | 50 | 2 | 0.9925 | 567 |
| LM | MH | 0.0361 | 0.0005 | 29961 | 827 | 51 | 1 | 0.9995 | 602 |
| GM |  | 0.0123 | 0.0000 | 68081 | 563 | 84 | 1 | 0.9997 | 575 |
| LM | TN | 0.0539 | 0.0010 | 6554 | 205 | 43 | 1 | 0.9988 | 532 |
| GM |  | 0.0186 | 0.0002 | 14110 | 282 | 64 | 1 | 0.9996 | 475 |

**Table 3.** Least square estimates and their respective standard errors of the parameters, *R*^2^ and AIC for the models fitted to the cumulative recovered cases of Covid-19 of the considered states.

| Model | State | b | SE | d | SE | e | SE | R2 | AIC |
| --- | --- | --- | --- | --- | --- | --- | --- | --- | --- |
| LM | AP | 0.0803 | 0.0011 | 1671975 | 605775 | 86 | 5.0819 | 0.9974 | 914 |
| GM |  | 0.0221 | 0.0004 | 1460882 | 125957 | 97 | 1.8518 | 0.9918 | 985 |
| LM | DL | 0.0882 | 0.0023 | 131453 | 1403 | 18 | 0.3475 | 0.9956 | 958 |
| GM |  | 0.0554 | 0.0011 | 142620 | 1343 | 13 | 0.2109 | 0.9984 | 906 |
| LM | KA | 0.0666 | 0.0010 | 2178476 | 904419 | 102 | 6.5403 | 0.9950 | 919 |
| GM |  | 0.0146 | 0.0002 | 2772194 | 1000 | 140 | 1.0527 | 0.9854 | 978 |
| LM | MH | 0.0351 | 0.0002 | 3284231 | 372300 | 116 | 3.6758 | 0.9985 | 984 |
| GM |  | 0.0106 | 0.0001 | 3728209 | 177040 | 139 | 1.2205 | 0.9943 | 1054 |
| LM | TN | 0.0540 | 0.0005 | 467663 | 11519 | 54 | 0.8245 | 0.9997 | 874 |
| GM |  | 0.0149 | 0.0001 | 1638075 | 22740 | 99 | 0.5812 | 0.9994 | 917 |

Our aim is not to analyze COVID-19 in all the states which have been considered. The aim of this paper is the analysis and prediction of the cumulative confirmed cases, cumulative deceased cases, and the cumulative recovered cases in Uttar Pradesh, India. However, we explored the situation of COVID-19 in other states of India to extract the prior information for the Bayesian non-linear regression models. Figures 2, 3 and 4 show the cumulative and the daily number of confirmed cases, deceased cases, and recovered cases of COVID-19 in Uttar Pradesh respectively and the fitted curve by the Baysian non-linear regression model using the prior information. In these figures, we see that due to the lack of randomness in cumulative confirmed cases, cumulative deceased cases, and cumulative recovered cases, the sum of the square residual is quite low.

**Figure 2.**
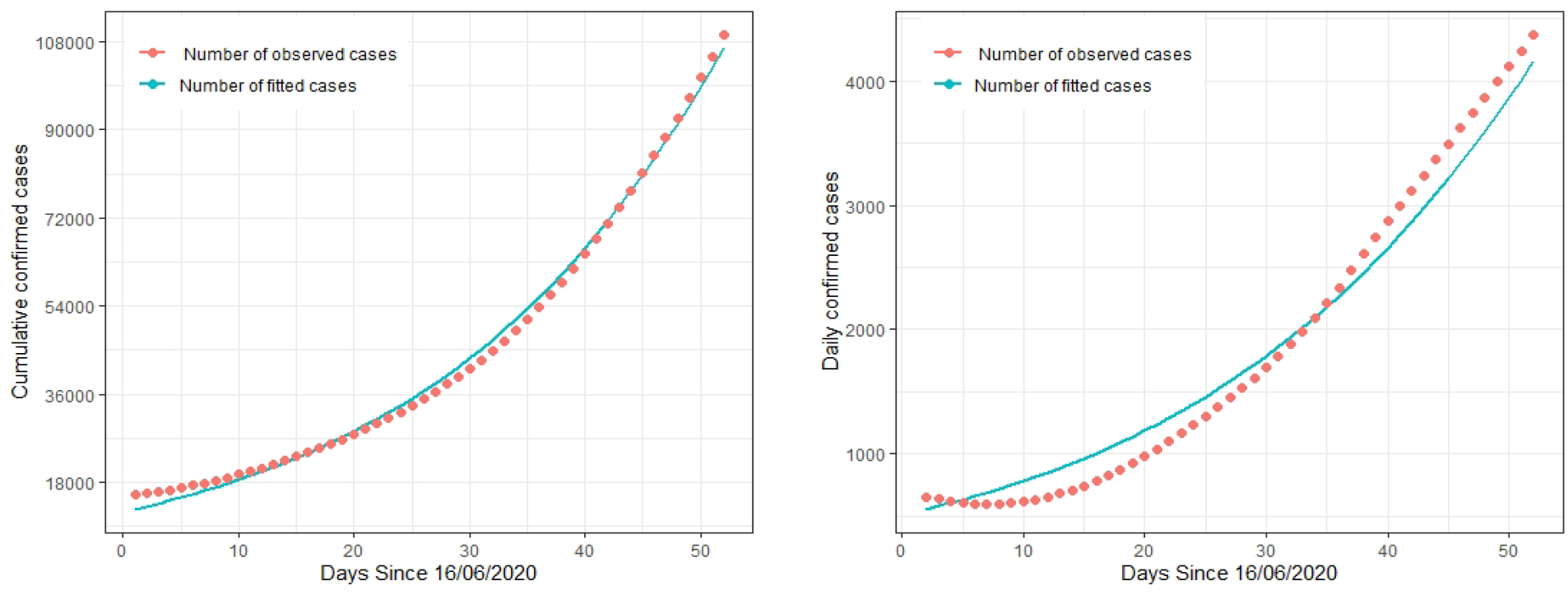
Cumulative and daily confirmed cases of Covid-19 in Uttar Pradesh, India, and fitted curve by Bayesian non-linear regression model using prior information.

**Figure 3.**
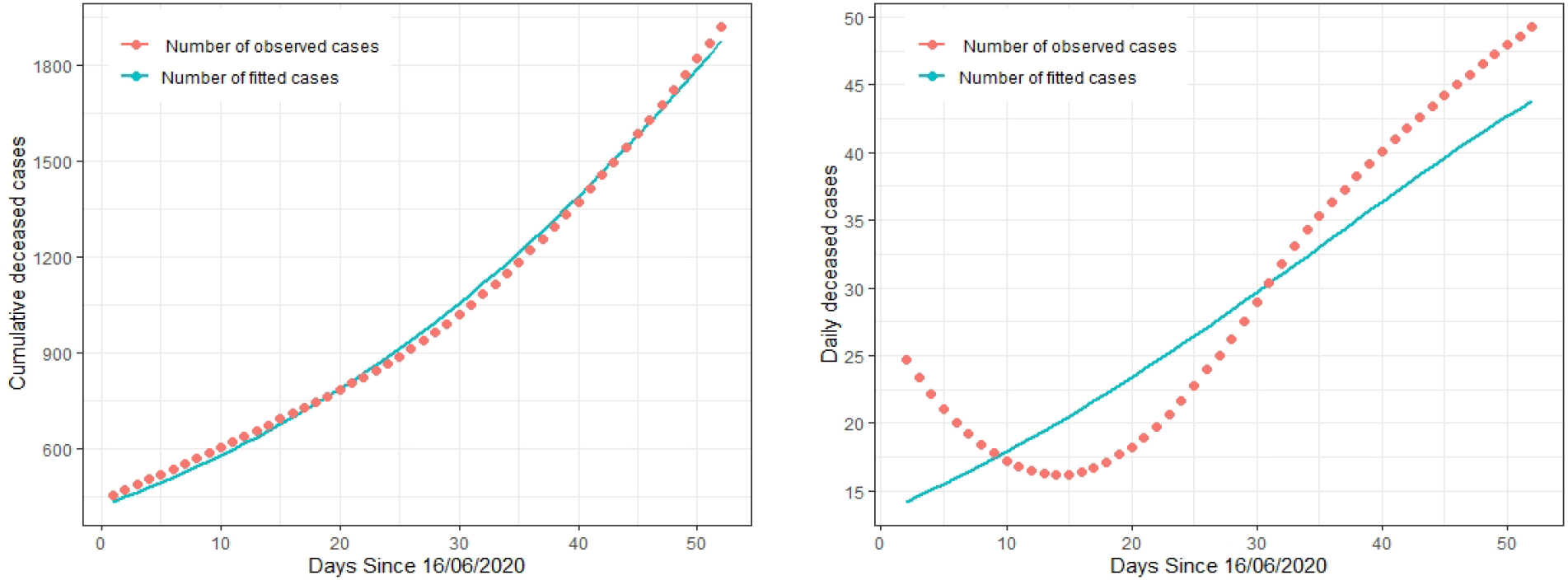
Cumulative and daily deceased cases of Covid-19 in Uttar Pradesh, India, and fitted curve by Bayesian non-linear regression model using prior information.

**Figure 4.**
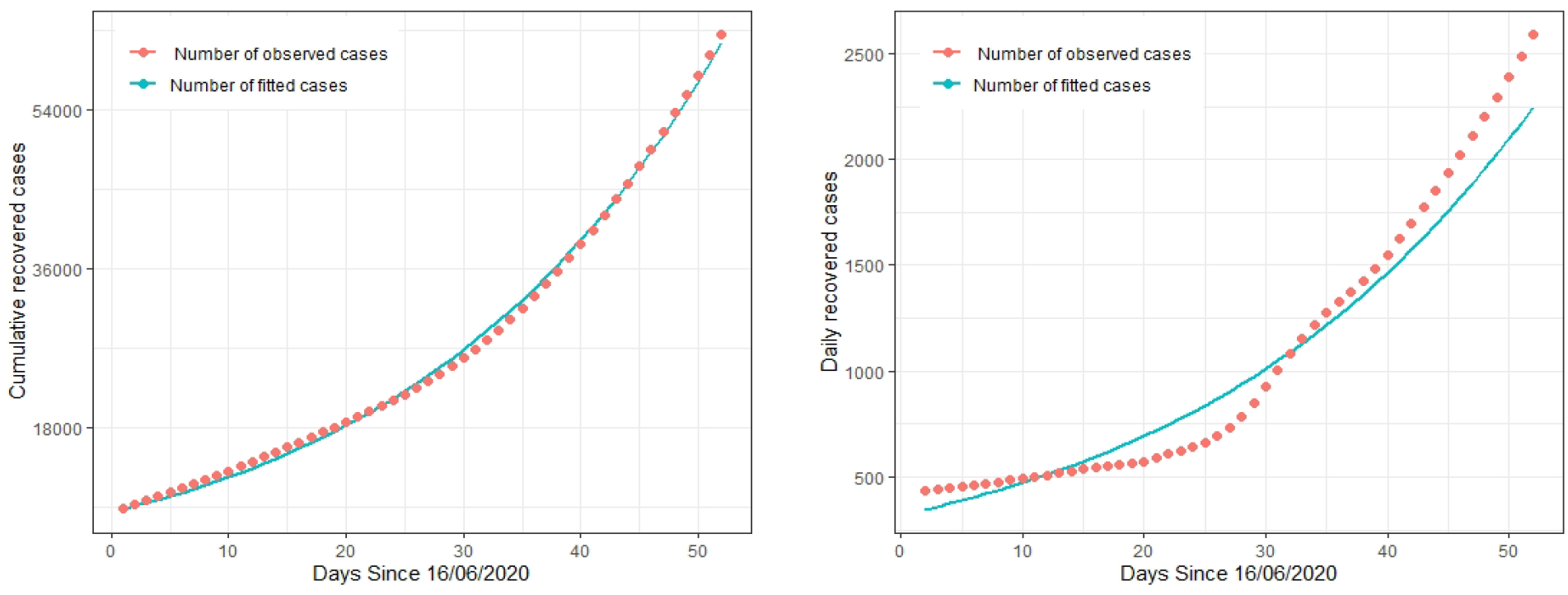
Cumulative and daily recovered cases of Covid-19 in Uttar Pradesh, India, and fitted curve by Bayesian non-linear regression model using prior information.

Tables 4, 5, and 6 represent the Bayesian estimates, standard error of the estimates, and *R*^2^ for the logistic and Gompertz models fitted to the cumulative confirmed cases, cumulative deceased cases and cumulative recovered cases of COVID-19 in UP respectively. Also, Watanabe Akaike information criterion (WAIC) is computed from the fitting of Bayesian Gompertz and logistic models to the data of the cumulative confirmed cases, cumulative deceased cases, and cumulative recovered cases of COVID-19 in UP, India. We used the prior distributions based on the least square estimates from the data of other considered states.

**Table 4.**
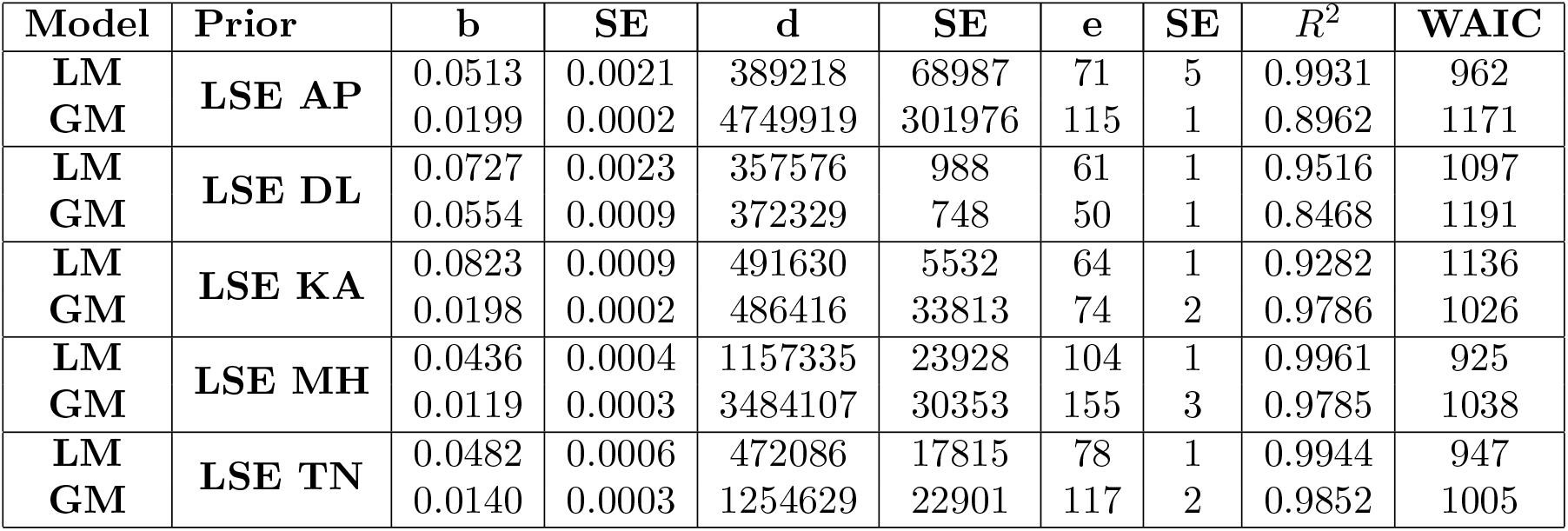
Bayesian estimates and their respective standard errors of the parameters, *R*^2^ and WAIC for the models fitted to the cumulative confirmed cases of Covid-19 in Uttar Pradesh, India by considering different prior distributions based on Least square estimated from data of other states of India.

**Table 5.**
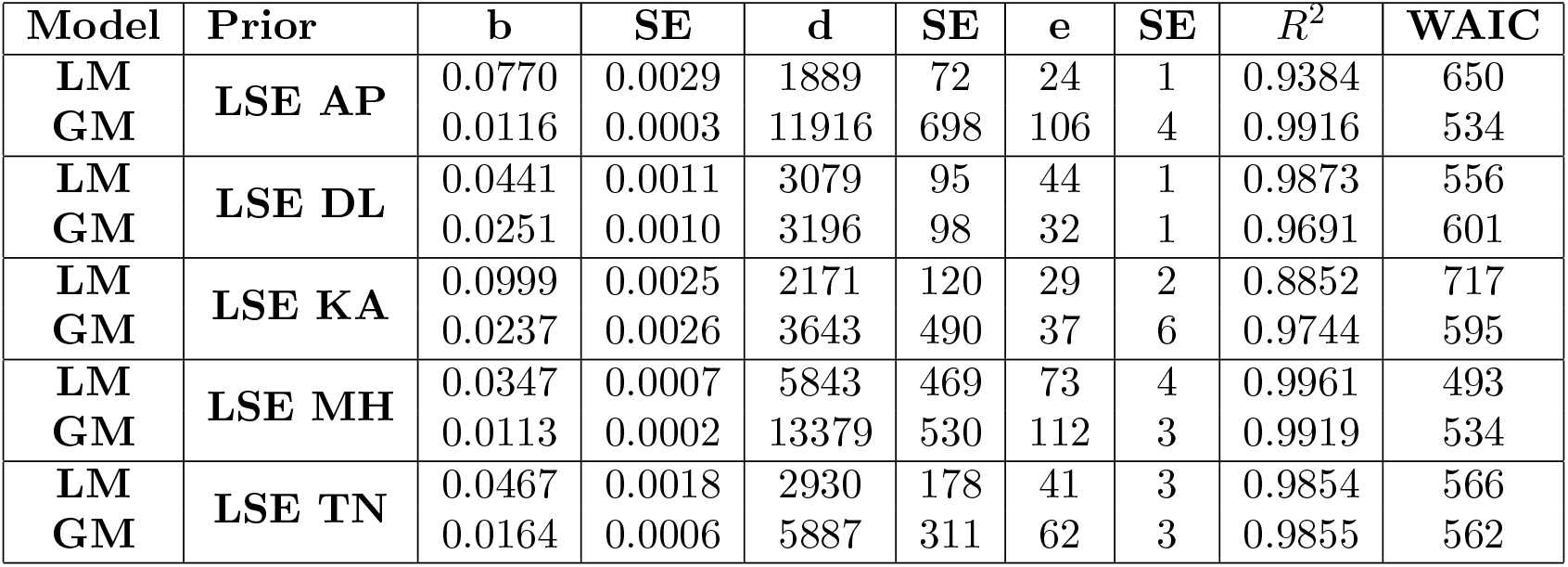
Bayesian estimates and their respective standard errors of the parameters, *R*^2^ and WAIC for the models fitted to the cumulative deceased cases of Covid-19 in Uttar Pradesh, India by considering different prior distributions based on Least square estimated from data of other states of India.

**Table 6.**
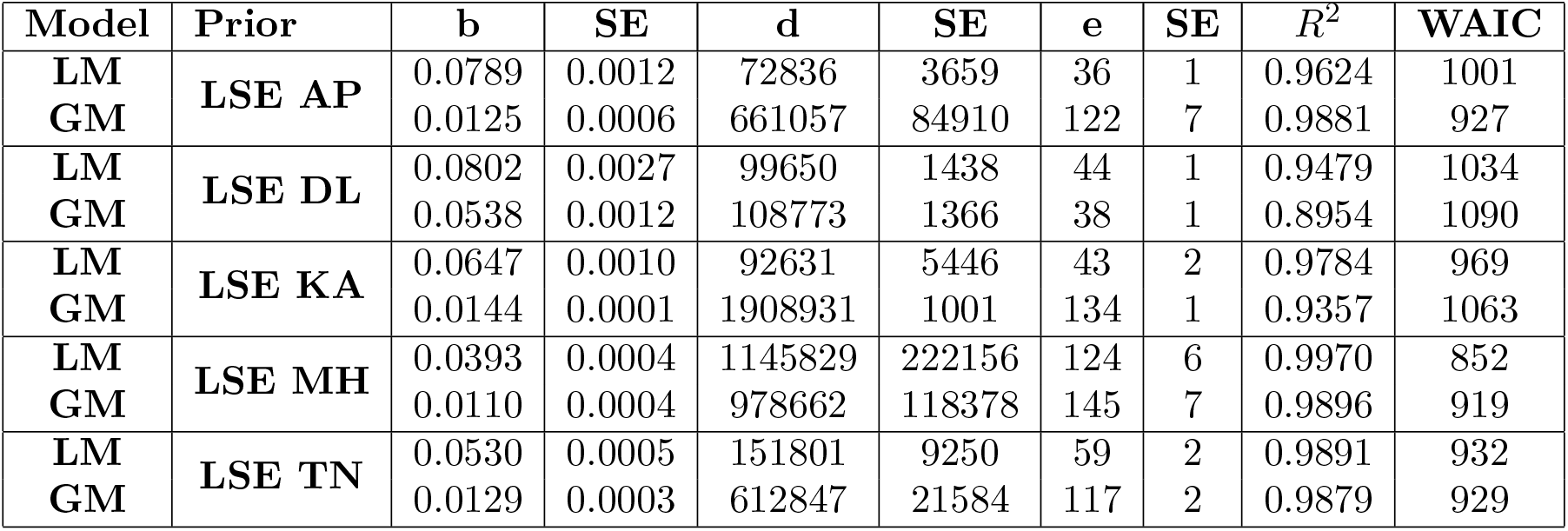
Bayesian estimates and their respective standard errors of the parameters, *R*^2^ and WAIC for the models fitted to the cumulative recovered cases of Covid-19 in Uttar Pradesh, India by considering different prior distributions based on Least square estimated from data of other states of India.

Based on the WAIC criterion, we can say that the COVID-19 pandemic curve of UP is quite closer to the Maharastra curve. This result does not conclude that the prevalence rate of both the state is equal, or they have equal cumulative confirmed cases, cumulative deceased cases, and cumulative recovered cases. The meaning of this result is that there is a similarity between the curve of both states. We used the least square estimates for Maharastra’s data as prior to predicting more accurate values. We can compare the situation of COVID-19 in both the states Maharastra and UP at the beginning of the pandemic. There were lackness of mass level screening of COVID-19 in both the states. Many people migrated from other countries to Mumbai, and there is no contact tracing in the beginning. Millions of people and labor class workers migrated from other states to UP, and there is a problem of contact tracing of the infected people.

In Figures 5, 6 and 7, we plotted the predicted curve by Bayesian non-linear regression model using prior information and the best fitted model, i.e., logistic model for UP state. The prediction results show that the model can predict the pandemic situation of COVID-19 very well for the considered cumulative and daily variables. Prediction is not well for the daily decreased number of cases, the possible explanation for this may be that the factors affecting the death rate are different from that of cumulative confirmed cases and cumulative recovered cases and hence, there is uncertainty in the daily number of deceased cases.

**Figure 5.**
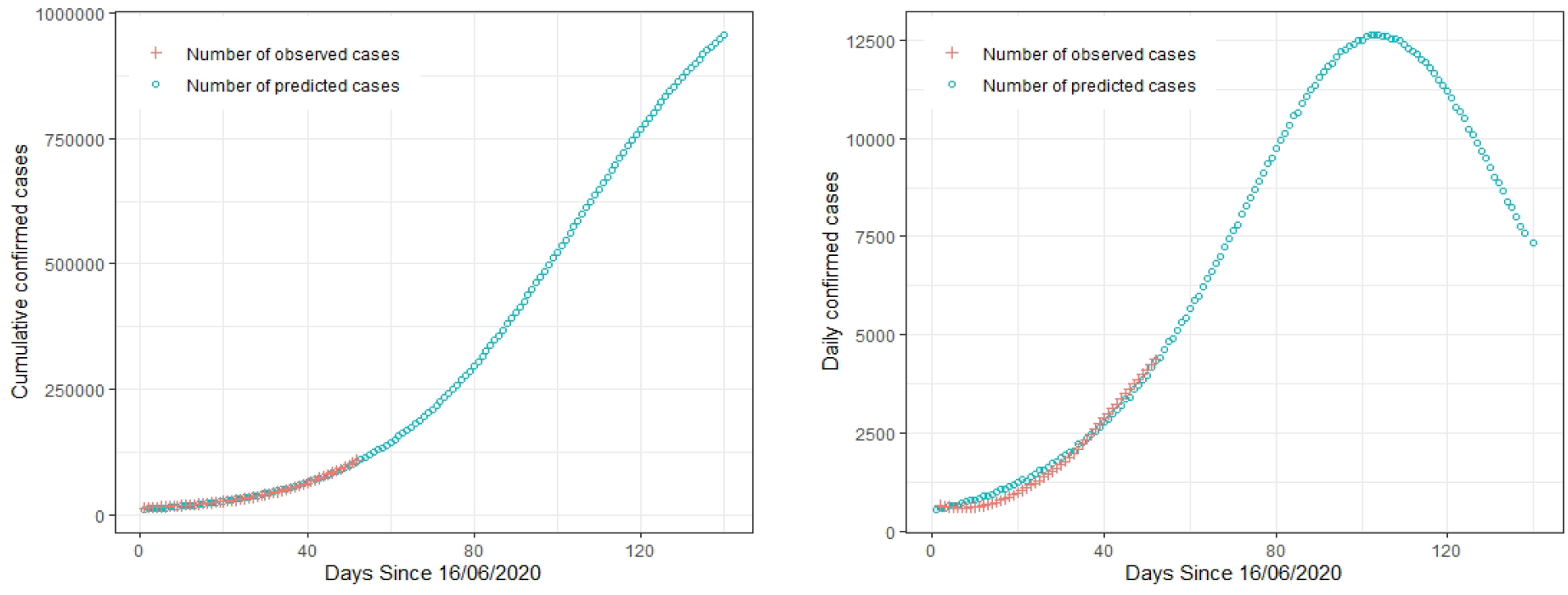
Cumulative and daily confirmed cases of Covid-19 in Uttar Pradesh, India, and predicted curve by Bayesian non-linear regression model using prior information.

**Figure 6.**
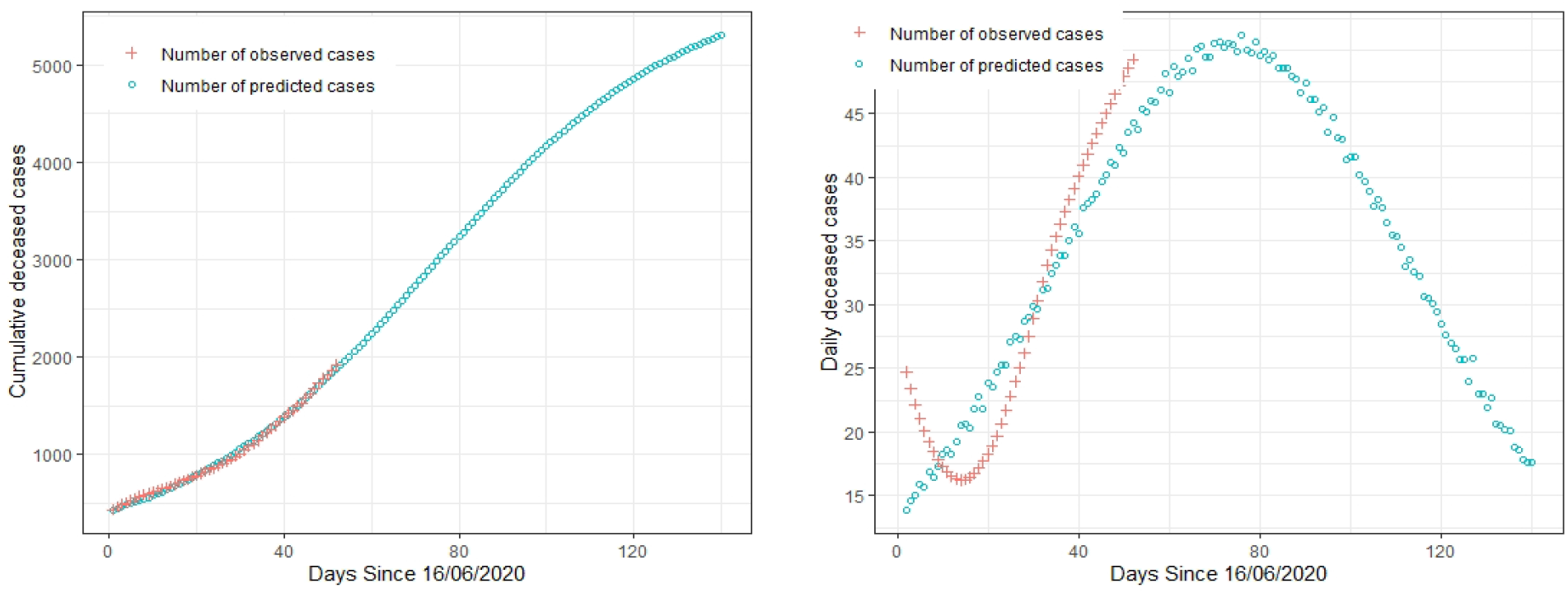
Cumulative and daily deceased cases of Covid-19 in Uttar Pradesh, India, and predicted curve by Bayesian non-linear regression model using prior information.

**Figure 7.**
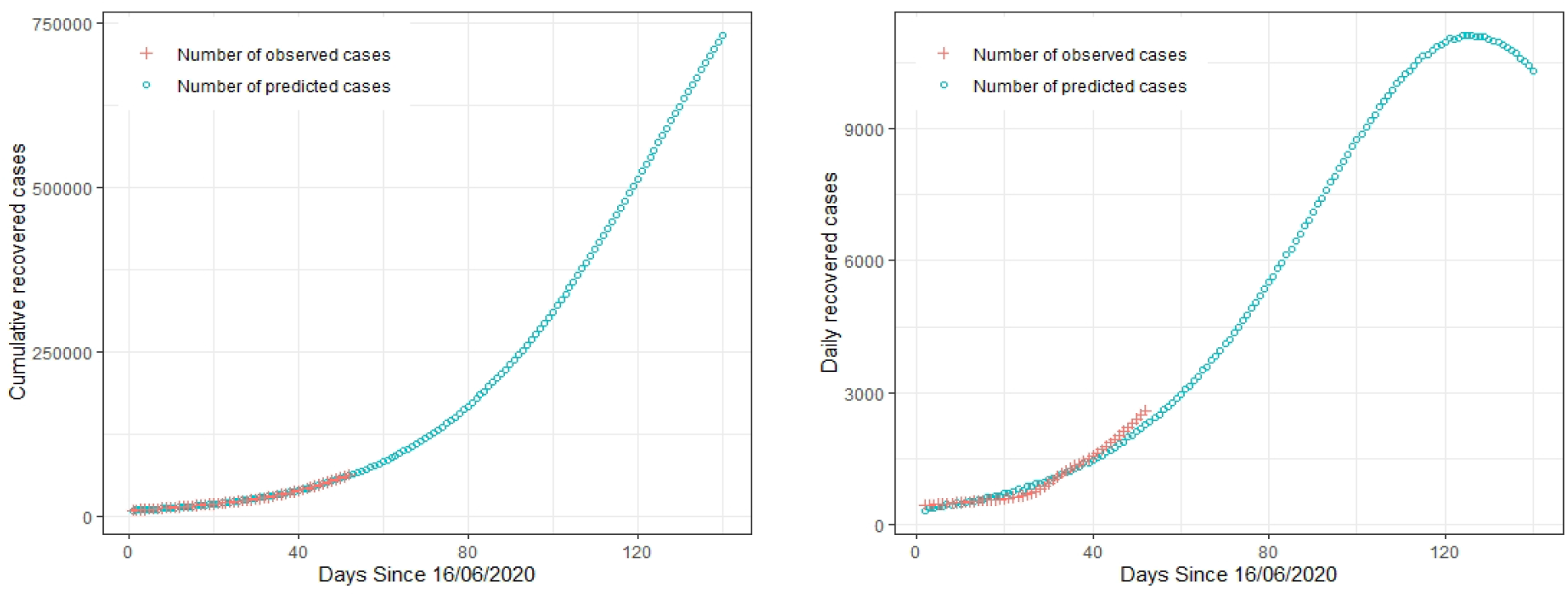
Cumulative and daily recovered cases of Covid-19 in Uttar Pradesh, India, and predicted curve by Bayesian non-linear regression model using prior information.

## 5. Conclusion

According to the fitting analysis of the existing data, the logistic model with the Maharashtra prior may be the best among the considered models studied in this paper. In Tables 7 and 8, we provided the predicted values of the daily and the cumulative number of confirmed cases, deceased cases and recovered cases of UP state. We presented the actual and predicted values up to 6 August 2020, and after that, only predicted values are given up to 25 September 2020. Here due to space constraints, we presented only five days interval and recent out of sample values at the daily level. From the tables, we conclude that the model predicts the values well to a great extent. Table 7 shows that the predicted daily confirmed cases on 25 September 2020 will be 12589, the predicted daily decreased number will be 40 on the same date, and the predicted daily recovered cases will be 9057 on the same date. Table 8 shows that the predicted cumulative confirmed cases on 25 September 2020 will be 548710, the predicted cumulative deceased number will be 4245 on the same date, and the predicted cumulative recovered cases will be 328662 on the same date.

**Table 7.** Prediction of daily confirmed, deceased and recovered cases of Covid-19 in Uttar Pradesh, India

| Days | Dates | Confirmed Cases |  | Deceased Cases |  | Recovered Cases |  |
| --- | --- | --- | --- | --- | --- | --- | --- |
|  |  | Daily Cases |  | Daily Cases |  | Daily Cases |  |
|  |  | Actual Cases | Prediction | Actual Cases | Prediction | Actual Cases | Prediction |
| 2 | 17/06/2020 | 583 | 593 | 30 | 15 | 335 | 357 |
| 7 | 22/06/2020 | 591 | 680 | 19 | 16 | 606 | 432 |
| 12 | 27/06/2020 | 606 | 820 | 19 | 19 | 632 | 508 |
| 17 | 02/07/2020 | 869 | 1076 | 17 | 20 | 592 | 645 |
| 22 | 07/07/2020 | 1332 | 1297 | 18 | 24 | 518 | 741 |
| 27 | 12/07/2020 | 1384 | 1635 | 21 | 28 | 645 | 907 |
| 32 | 17/07/2020 | 1722 | 1897 | 38 | 31 | 959 | 1091 |
| 37 | 22/07/2020 | 2300 | 2391 | 34 | 34 | 1645 | 1307 |
| 42 | 27/07/2020 | 3505 | 2862 | 30 | 39 | 1192 | 1567 |
| 43 | 28/07/2020 | 3458 | 2982 | 41 | 38 | 1687 | 1638 |
| 44 | 29/07/2020 | 3383 | 3102 | 33 | 38 | 1287 | 1697 |
| 45 | 30/07/2020 | 3705 | 3238 | 57 | 40 | 996 | 1738 |
| 46 | 31/07/2020 | 4422 | 3293 | 43 | 41 | 2060 | 1821 |
| 47 | 01/08/2020 | 3587 | 3480 | 47 | 40 | 2471 | 1892 |
| 48 | 02/08/2020 | 3873 | 3612 | 53 | 41 | 2023 | 1961 |
| 49 | 03/08/2020 | 4441 | 3686 | 48 | 43 | 2036 | 2012 |
| 50 | 04/08/2020 | 2948 | 3889 | 39 | 42 | 1878 | 2101 |
| 51 | 05/08/2020 | 4078 | 4035 | 40 | 44 | 3287 | 2154 |
| 52 | 06/08/2020 | 4586 | 4115 | 61 | 44 | 2844 | 2278 |
| 57 | 11/08/2020 |  | 4950 |  | 46 |  | 2684 |
| 62 | 16/08/2020 |  | 5816 |  | 49 |  | 3166 |
| 67 | 21/08/2020 |  | 6820 |  | 49 |  | 3723 |
| 72 | 26/08/2020 |  | 7861 |  | 50 |  | 4326 |
| 77 | 31/08/2020 |  | 8911 |  | 50 |  | 5049 |
| 82 | 05/09/2020 |  | 9919 |  | 49 |  | 5809 |
| 87 | 10/09/2020 |  | 10857 |  | 49 |  | 6616 |
| 92 | 15/09/2020 |  | 11689 |  | 45 |  | 7443 |
| 97 | 20/09/2020 |  | 12296 |  | 43 |  | 8273 |
| 102 | 25/09/2020 |  | 12589 |  | 40 |  | 9057 |

**Table 8.** Prediction of cumulative confirmed, deceased and recovered cases of Covid-19 in Uttar Pradesh, India

|  |  | Confirmed Cases |  | Deceased Cases |  | Recovered Cases |  |
| --- | --- | --- | --- | --- | --- | --- | --- |
|  |  | Cumulative Cases |  | Cumulative Cases |  | Cumulative Cases |  |
| Days | Dates | Actual Cases | Prediction | Actual Cases | Prediction | Actual Cases | Prediction |
| 2 | 17/06/2020 | 15181 | 13170 | 465 | 451 | 9239 | 9058 |
| 7 | 22/06/2020 | 18322 | 16273 | 569 | 528 | 11601 | 11011 |
| 12 | 27/06/2020 | 21549 | 20180 | 649 | 618 | 14215 | 13370 |
| 17 | 02/07/2020 | 24825 | 25043 | 735 | 719 | 17221 | 16267 |
| 22 | 07/07/2020 | 29968 | 30962 | 827 | 837 | 19627 | 19699 |
| 27 | 12/07/2020 | 36476 | 38265 | 934 | 969 | 23334 | 23886 |
| 32 | 17/07/2020 | 45163 | 47143 | 1084 | 1117 | 27634 | 28946 |
| 37 | 22/07/2020 | 55588 | 58099 | 1263 | 1282 | 33500 | 35038 |
| 42 | 27/07/2020 | 70493 | 71407 | 1456 | 1465 | 42833 | 42361 |
| 43 | 28/07/2020 | 73951 | 74389 | 1497 | 1503 | 44520 | 43999 |
| 44 | 29/07/2020 | 77334 | 77491 | 1530 | 1541 | 45807 | 45696 |
| 45 | 30/07/2020 | 81039 | 80729 | 1587 | 1581 | 46803 | 47434 |
| 46 | 31/07/2020 | 85461 | 84022 | 1630 | 1622 | 48863 | 49255 |
| 47 | 01/08/2020 | 89048 | 87502 | 1677 | 1662 | 51334 | 51147 |
| 48 | 02/08/2020 | 92921 | 91114 | 1730 | 1703 | 53357 | 53108 |
| 49 | 03/08/2020 | 97362 | 94800 | 1778 | 1746 | 55393 | 55120 |
| 50 | 04/08/2020 | 100310 | 98689 | 1817 | 1788 | 57271 | 57221 |
| 51 | 05/08/2020 | 104388 | 102724 | 1857 | 1832 | 60558 | 59375 |
| 52 | 06/08/2020 | 108974 | 106839 | 1918 | 1876 | 63402 | 61653 |
| 57 | 11/08/2020 |  | 129962 |  | 2102 |  | 74156 |
| 62 | 16/08/2020 |  | 157357 |  | 2340 |  | 88959 |
| 67 | 21/08/2020 |  | 189459 |  | 2586 |  | 106440 |
| 72 | 26/08/2020 |  | 226621 |  | 2837 |  | 126893 |
| 77 | 31/08/2020 |  | 269030 |  | 3089 |  | 150759 |
| 82 | 05/09/2020 |  | 316645 |  | 3338 |  | 178270 |
| 87 | 10/09/2020 |  | 369271 |  | 3582 |  | 209711 |
| 92 | 15/09/2020 |  | 426133 |  | 3815 |  | 245238 |
| 97 | 20/09/2020 |  | 486372 |  | 4037 |  | 284926 |
| 102 | 25/09/2020 |  | 548710 |  | 4245 |  | 328662 |

The predicted maximum cumulative number of confirmed cases, deceased cases, and recovered cases by all the considered models will be: 357576-4749919, 1889-11916, and 72836-1908931 respectively as per the current trend. The daily number of confirmed, deceased, and recovered cases will be maximum in between 50-155 days, 24-112 days and 36-145 days from 16 June 2020 respectively. In Table 9, we provided the predicted maximum cumulative number of confirmed cases, deceased cases and recovered cases and the predicted date when theses cases reach to the maximum using the best-fitted model in UP state. Results show that the maximum number of cumulative number of confirmed cases will be 1157335 on 3 June 2021. The maximum number of cumulative deceased will reach to the 5843 on 28 March 2021 and the maximum number of recovered cases will be 1145829 on 1 July 2021. The daily number of confirmed, deceased and recovered cases will be maximum at 104th day, 73rd day and 124th day from 16 June 2020 using the best-fitted model. The results of the best fitted model also show that the COVID-19 will be over probably by early-June, 2021. We conclude that the proposed method can be useful, and we believe this study can provide some valuable information to strengthen the implementation of strategies to increase the health system capacity and also help the public health authorities to make the relevant decision.

**Table 9.** Maximum predicted number of cumulative confirmed, deceased and recovered cases of Covid-19 in Uttar Pradesh, India and the predicted date when these cases reach to the maximum

| Cases | Predicted cumulative number of cases | Predicted date when the cumulative number of cases reach to the maximum |
| --- | --- | --- |
| Confirmed Cases | 1157335 | 03/06/2021 |
| Deceased Cases | 5843 | 28/03/2021 |
| Recovered Cases | 1145829 | 01/07/2021 |

## Data Availability

In this paper, we used the following major databases:
1. Github-Covid-19 in India available from https://github.com/imdevskp/covid-19-india-data
2. Covid19India is a open source database for India available from: https://www.covid19india.org/

https://www.covid19india.org/

https://github.com/imdevskp/covid-19-india-data

